# Prevalence and risk factors of low back pain among nursing students

**DOI:** 10.1101/2023.11.15.23298588

**Authors:** Jin Liya, Zhu Ya, Hu Miaoye, Zhang Chunmei

## Abstract

**Study design:** Retrospective case-control study

**Objective:** To investigate the incidence of low back pain among nursing students and to analyze the factors contributing to low back pain among nursing students.

**Method:** This retrospective study was conducted in 78 nursing students who entered our hospital for internship from July 2022 to March 2023. Demographic data, including age, gender, height, weight, exercise habits, staying-up-late habits, smoking history, education level, mental status, working posture, menstrual pain, and other basic information were collected, and the number of interns experiencing low back pain in the past 1 month and 3 months was recorded to investigate the incidence of low back pain among nursing students. Relevant risk factors were also analyzed.

**Results:** The incidence of low back pain among nursing students was 19.2% within 1 month and 25.6% within 3 months. Factors such as gender, age, height, weight, smoking history, exercise habits, staying-up-late habits, education level, mental status, working posture correctness, and menstrual pain showed no statistical significance between the pain and non-pain groups (*P* > 0.05), while BMI and exercise habits showed statistical differences between the two groups (*P* < 0.05). BMI (*OR* = 1.530; 95% confidence interval = 1.16 - 2.02; *P* = 0.003) was identified as a risk factor for low back pain. Exercise habits (*OR* = 0.232; 95% confidence interval = 0.07 - 0.81; *P* = 0.022) were identified as protective factors for low back pain.

**Conclusion:** The incidence of low back pain among nursing students remains relatively high, and low back pain is associated with high BMI and lack of exercise. These results can provide a reference for developing preventive measures for low back pain among nursing students.

## Introduction

Pain is an unpleasant emotional state and serves as a defense mechanism to protect injured parts of the body from further harm. Low back pain, more accurately termed as lumbosacral pain, occurs below the 12th rib and above the buttocks. Work-related issues in the lower back are highly prevalent among professional nurses, who are considered one of the professional groups in the healthcare service system. A review of 132 research papers on musculoskeletal diseases related to nursing work indicated that the annual average prevalence of low back pain among nurses is 55% [1]. Low back pain often leads to restricted work and life activities or disability, posing a threat to the productivity and stability of the registered nurse workforce [2-4].

Occupational nurses’ low back pain ranks third among musculoskeletal health issues in this group. This issue is associated with the physical activities and ergonomics risk factors in the workplace for nurses [5]. On one hand, nurses frequently engage in high-intensity physical activities, such as manual lifting or patient handling [6], which often lead to injuries in the lower back muscles, ligaments, and other soft tissues. On the other hand, nurses lack awareness of low back pain prevention and related training, making them susceptible to low back pain, thereby increasing absenteeism and early retirement due to poor health conditions among nurses [7, 8]. Moreover, the probability of low back pain increases with extended working hours and reduced rest time, accumulating over time [9]. Nursing students need to complete relevant skill training during clinical rotations, thus facing similar occupational exposures. Due to their lack of nursing-related work experience and awareness of low back care, nursing students are more prone to developing low back pain [10]. Furthermore, most nursing schools currently lack relevant strategies for preventing and training nursing students in low back pain protection. Once nursing students experience low back pain, it not only affects their physical well-being in daily life and work but also leads to psychological issues such as aversion to the nursing profession and disinterest in learning [11, 12]. Although there has been extensive research in recent years on the prevalence, influencing factors, and interventions for low back pain among registered nurses [13, 14], a review of literature has not found relevant studies on low back pain among nursing students in Mainland China. In this study, we collected relevant data from nursing students who entered our hospital for internship and analyzed the factors influencing low back pain.

## Materials and Methods

### Study Population

A retrospective study was conducted on nursing students who entered our hospital for internship from July 2021 to March 2022. Inclusion criteria: completion of university courses with passing grades, good physical health, emotional self-control, and voluntary participation. Exclusion criteria: absence from training for more than 1 week due to leave, travel, or other reasons, inability to complete training and withdrawal midway, presence of musculoskeletal diseases, and underlying medical conditions. A total of 78 nursing students were included in the study. Data on age, gender, height, weight, exercise habits (defined as having ≥ 2 sessions of ≥ 30 minutes of exercise per week, otherwise considered as no exercise habit), staying-up-late habits (defined as staying up past 1 am for ≥ 4 nights per week, otherwise considered as no staying-up-late habit), smoking history, education level, mental status (whether anxiety-prone in daily life), correctness of working posture, and menstrual pain in females were collected. The number of interns experiencing low back pain in the past 1 month and 3 months was recorded. The diagnostic criteria for low back pain in this study were as follows: feeling of soreness, swelling, pain, or discomfort in the lower back, relief after rest and aggravation after fatigue or activity; relief of symptoms with a supporting pad under the lower back when lying down, inability to bend or worsening of pain symptoms; no abnormality in the shape of the lower back, localized tenderness, and negative findings on imaging studies. The visual analogue scale (VAS) was used to score low back pain, with a score > 3 indicating moderate or severe pain lasting at least one day. Data from nursing students who had and had not experienced low back pain were compared to analyze the factors contributing to low back pain.

### Data Collection

The survey was conducted in the form of a questionnaire. After standardized training, surveyors distributed the questionnaires to the interns with their consent, and the interns filled in the questionnaire with their real names. The completion time for the survey did not exceed 20 minutes, and the completed questionnaires were collected on-site. All questionnaire data were verified by the surveyors and summarized and organized using WPS spreadsheets.

### Statistical Analysis

All data were analyzed using SPSS 26.0 (SPSS, Chicago, Illinois, USA). T-tests were used to compare continuous variables, and the Pearson’s chi-squared test and the continuity-corrected chi-squared test were used to compare categorical variable data, with a significance level set at *P* < 0.05. For independent risk factors, binary logistic regression analysis was performed. Variables with significant differences (*P* < 0.05) were included in the final binary logistic regression analysis. Variables that did not meet *P* < 0.05 were removed, and the results were reported with a 95% confidence interval. Statistical significance was defined as *P* < 0.05.

## Results

### Low Back Pain Status among Nursing Students

This retrospective study included a total of 78 nursing students. The demographic characteristics and relevant data of all nursing students are presented in Table 1. Among them, there were 8 male nursing students (10.3%) and 70 female nursing students (89.7%). The average age of all nursing students was 22.2 ± 0.85 years, with an average height of 162.1 ± 4.63 cm, average weight of 50.2 ± 6.12 kg, and an average BMI of 21.8 ± 1.21 kg/m^2^. The number of interns experiencing low back pain within 1 month was 15, accounting for 19.2%, and within 3 months was 20, accounting for 25.6%. Detailed statistics on other survey factors are presented in Table 1.

**Table 1.** Demographic Characteristics and and Prevalence of Back Pain in Nursing Students.

| Items | N (%) |
| --- | --- |
| Gender |  |
| male | 8 (10.3%) |
| female | 70 (89.7%) |
| Age/y, $x \pm s$ | $22.2 \pm 0.85$ |
| Height/cm, $x \pm s$ | $162.1 \pm 4.63$ |
| Weight/kg, $x \pm s$ | $50.2 \pm 6.12$ |
| BMI, $x \pm s$ | $21.8 \pm 1.21$ |
| Exercise habits | 39 (50.0%) |
| Smoking history | 4 (5.1%) |
| Staying-up-late habits | 31 (39.7%) |
| Education level (bachelor degree or above) | 60 (76.9%) |
| Mental status (prone to anxiety) | 11 (14.1%) |
| Female menstrual pain | 34 (48.5%) |
| Working posture correctness | 7 (9.9%) |
| Low back pain within 1 month | 15 (19.2%) |
| Low back pain within 3 month | 20 (25.6%) |

### Risk Factors for Low Back Pain among Nursing Students

Nursing students were categorized into non-pain group and pain group based on whether they experienced continuous pain, soreness, or numbness in the lower back for at least one day in the past 1 month and 3 months, as shown in Table 2. There were no statistical differences (*P* > 0.05) between the non-pain group and the pain group in terms of gender, age, height, weight, smoking history, staying-up-late habits, education level, mental status, correctness of working posture, and menstrual pain in females. However, there were statistical differences (*P* < 0.05) in terms of BMI and exercise habits. Through binary logistic regression analysis (see Table 3), we found that BMI (*OR =* 1.530; 95% confidence interval = 1.16 - 2.02; *P* = 0.003) was identified as a risk factor for low back pain. Additionally, exercise habits (*OR* = 0.232; 95% confidence interval = 0.07 - 0.81; *P* = 0.022) were identified as protective factors for low back pain.

**Table 2.** Univariate Analysis of Risk Factors for Back Pain in Nursing Students.

| Group | No pain | Pain | P |
| --- | --- | --- | --- |
| Gender |  |  | 0.420 |
| male | 5 | 3 |  |
| female | 53 | 17 |  |
| Age/y, $x \pm s$ | $22.3 \pm 0.87$ | $21.9 \pm 0.83$ | 0.532 |
| Height/cm, $x \pm s$ | $161.7 \pm 4.28$ | $163.4 \pm 4.68$ | 0.139 |
| Weight/kg, $x \pm s$ | $48.9 \pm 6.67$ | $51.8 \pm 6.88$ | 0.102 |
| BMI, $x \pm s$ | $21.2 \pm 1.29$ | $23.7 \pm 1.47$ | <0.01 |
| Exercise habits |  |  | 0.039 |
| No | 25 | 14 |  |
| Yes | 33 | 6 |  |
| Smoking history |  |  | 0.976 |
| No | 55 | 19 |  |
| Yes | 3 | 1 |  |
| Staying-up-late habits |  |  | 0.108 |
| No | 38 | 9 |  |
| Yes | 20 | 11 |  |
| Bachelor degree or above |  |  | 0.814 |
| No | 13 | 5 |  |
| Yes | 45 | 15 |  |
| Prone to anxiety |  |  | 0.383 |
| No | 51 | 16 |  |
| Yes | 7 | 4 |  |
| Female menstrual pain |  |  | 0.887 |
| No | 27 | 9 |  |
| Yes | 26 | 8 |  |
| Working posture correctness |  |  | 0.852 |
| No | 53 | 18 |  |
| Yes | 5 | 2 |  |

**Table 3.** Multivariate Analysis of Independent Risk Factors for Back Pain in Nursing Students.

| Variable | *P* | *OR* | 95% *CI* |
| --- | --- | --- | --- |
| BMI | 0.003 | 1.530 | 1.16-2.02 |
| Exercise habits | 0.022 | 0.232 | 0.07-0.81 |

## Discussion

Despite numerous studies on the back problems of nursing staff, most researches have focused on registered nurses, with only a few studies investigating nursing students [15, 16]. A search of literature in Mainland China has not yet revealed studies on back pain among nursing students. Smith and Leggat [15] found a relatively high prevalence of back pain among nursing students, suggesting that nursing managers also need to pay attention to the issue of back pain among nursing students. In our study, we also found that some nursing students had already experienced back pain upon entering our hospital for internship, with a three-month cumulative incidence of back pain as high as 25.6%. This may be due to the daily learning and skills training in nursing school. Similarly, the lack of cognitive and protective training during the internship may increase the likelihood of developing back pain when nursing students participate in clinical nursing work. The occurrence of back pain during the internship may have an impact on the career choices of nursing students, and may even lead to dropping out of the program. However, clinical preparation is an important part of nursing education, and clinical training is its foundation. Work-related musculoskeletal disorders are important occupational health issues in nursing, and research in recent years has been increasing [17]. Therefore, it is important to identify the risk factors that may lead to back pain in nursing students and take necessary intervention measures to protect them. Nurses play an important role in promoting the health of patients, but their own health is the prerequisite for ensuring the profession. Therefore, nursing students should be encouraged to pay attention to their physical health and take necessary protective measures to maintain their own health [19].

In this study, we found that BMI is a risk factor for back pain. With increasing BMI, excess fat around the waist and hips can cause pelvic tilt, leading to abnormal mechanical relationships between the hip flexors and extensors. The hip flexors become tighter, while the hip extensors become longer and weaker, increasing the probability of back pain. Additionally, an increase in BMI increases the mechanical pressure on the thoracolumbar spine and paraspinal muscles, leading to higher pressure on the muscles of the lower back in daily life and work. Furthermore, if BMI reaches the overweight or obese standard, muscles are prone to inflammation in this state, affecting the nutritional blood vessels of the musculoskeletal system, leading to degeneration and pain in the bones and muscles. Zeng et al. [20] found in their study that BMI is a risk factor for back pain among registered nurses. They also found that factors such as years of work experience, number of night shifts, and psychological fatigue have a certain impact on the occurrence of back pain. Similarly, Wu et al. [21] confirmed that BMI is associated with a higher incidence of back pain among registered nurses. However, their study found that the shorter the working years, the higher the probability of back pain, which may be related to the lack of clinical experience.

The lower back muscles, as one of the core muscle groups, play an important role in maintaining the balance and stability of the trunk. The anatomical structure of the lower back determines that it will bear additional abnormal loads when sitting for a long time, bending over, lifting heavy objects, and adopting incorrect working postures, leading to lower back pain or even lumbar degeneration over time. The nature of the work, especially those working in departments such as the ICU, determines that the probability of back pain will be greatly increased. Many studies [22, 23] have confirmed this, but this study did not find this factor to be a risk factor for back pain, possibly because the samples included in this study were nursing students who had just entered or had not yet entered clinical rotations, and the rate of poor posture was lower. Prior to intervention, both groups had received correct posture training for related lifting, carrying, and bending operations. The workload assigned to nursing students was relatively light. In addition, factors such as smoking, staying up late, mental state, and education level, which are risk factors for back pain in some practicing nurses, were not found to be risk factors for back pain among nursing students in this study. This may be related to the age and living environment of the students. Therefore, it is possible that the risk factors for back pain among nursing students in different internship stages may also be different, which requires further research to clarify.

The protective factor of exercise habits, confirmed in this study, has been rarely mentioned in previous reports. Exercise provides a certain degree of training for the body’s functions and related musculoskeletal muscles, enabling nursing students to better cope with various physically demanding tasks during clinical nursing work. Similarly, exercise can increase limb coordination and improve back pain caused by poor posture. Many studies have found that nursing students who undergo training in back muscle exercises have significantly reduced back pain. Tian et al. [9] designed back muscle exercise interventions for ICU nurses and confirmed that back muscle exercises can alleviate back pain in professional nurses. Bai et al. [22] found that the application of certain yoga movements in ICU nurses can also alleviate back pain. However, providing only a back muscle exercise manual and clinical nursing experience teaching did not lead to a significant improvement in back pain among nursing students, which may be related to the need to improve compliance with back muscle exercise in nursing students. In the study by Lin et al. [23], it was found that providing nurses with videos about back pain prevention was more effective in improving compliance than distributing manuals, reflecting compliance issues. In routine clinical nursing work, compliance can be improved by implementing a reasonable scheduling system to enhance flexibility. When implementing intervention measures for nursing students, attention should be paid to the safety of the intervention measures, avoiding excessive training, and carrying out back muscle exercises under the guidance of the training and teaching team, in order to improve the physical and mental health of nursing students, enhance their quality of life, and have a positive impact on their clinical nursing work and life after formally entering the medical and health service system.

In this study, we found that among the 78 nursing students included in the study, 15 people experienced pain within one month, accounting for 19.2%, while 20 people experienced pain within three months, accounting for 25.6%. Previous studies [15, 24] have also investigated the prevalence of back problems among nursing students. A prevalence study conducted by Smith and Leggat [15] even found that the annual prevalence of back pain among nursing students in rural areas of Australia (59.2%) was higher than that of some registered nurses. Menzel et al. [25] reported an annual prevalence of work-related back pain among nursing staff of 36.9%-75%. Our study found that the back problems of nursing students began during the school training period and were exacerbated during the internship. This suggests that the training courses in nursing school need to be adjusted to some extent, or interventions during school may be preferable. Research has shown that exercise intervention measures can alleviate the accumulated muscle fatigue of nursing students after participating in clinical nursing work, reduce muscle inflammation, and thus reduce the occurrence of back pain. At the same time, exercise strengthens the back muscles and improves the coordination of the back muscles, enabling nursing students to adapt to daily work and maintain an awareness of protecting the back muscles. However, relying solely on back muscle exercises is not enough to completely solve the problem of back pain among nursing students. The back problems of nursing students are the result of multiple factors, including physiological factors such as BMI, as well as psychological factors and work environment factors. Therefore, it is necessary to implement relevant intervention measures for nursing students from different levels and perspectives, which depends on the establishment of a comprehensive intervention system for back pain among nursing students in the medical and health service system and medical education system, in order to reduce the occurrence of back pain among nursing students and enhance their ability to protect against back pain.

## Data Availability

All data produced in the present study are available upon reasonable request to the authors

